# Detection of neutralizing antibodies against SARS-CoV-2 by using a commercial surrogate virus neutralization ELISA: can it substitute the classical neutralization test?

**DOI:** 10.1101/2021.10.12.21264881

**Authors:** Natalie Hofmann, Marica Grossegesse, Markus Neumann, Lars Schaade, Andreas Nitsche

## Abstract

**Background:** High-throughput detection of neutralizing antibodies against SARS-CoV-2 presents a valuable tool for vaccine trials or investigations of population immunity. We evaluate the performance of the first commercial surrogate virus neutralization test (sVNT, GenScript Biotech) against SARS-CoV-2 plaque reduction neutralization test (PRNT) in convalescent and vaccinated individuals. We compare it to five other ELISAs, two of which are designed to detect neutralizing antibodies.

**Results:** In 491 pre-vaccination serum samples, sVNT missed 23.6% of PRNT-positive samples when using the manufacturer-recommended cutoff of 30% binding inhibition. Introducing a equivocal area between 15 and 35% maximized sensitivity and specificity against PRNT to 72.8–93.1 % and 73.5– 97.6%, respectively. The overall diagnostic performance of the other ELISAs for neutralizing antibodies was below that of sVNT. Vaccinated individuals exhibited higher antibody titers by PRNT (median 119.8, IQR 56.7–160) and binding inhibition by sVNT (median 95.7, IQR 88.1–96.8) than convalescent patients (median 49.1, IQR 20–62; median 52.9, IQR 31.2–76.2).

**Conclusion:** GenScript sVNT is suitable to screen for SARS-CoV-2-neutralizing antibodies; however, to obtain accurate results, confirmatory testing by PRNT in a equivocal area is required. This equivocal area may require adaptation for use in vaccinated individuals, due to higher antibody titers.

## Introduction

Over the course of the COVID-19 pandemic and especially with the availability of several vaccines, the detection of neutralizing antibodies as a potential marker of immunity has become increasingly important for use in vaccine trials, to establish individual vaccination success or to evaluate the population immunity to infection and disease. While it is still under discussion whether there is a certain antibody titer that confers protection to SARS-CoV-2 (correlate of protection, CoP), as is the case for other viruses – for example, for Hepatitis B virus an antibody titer above 10 mIU/mL is associated with protection against infection [1] –, recent studies suggest that neutralizing antibodies present a good estimate for protection also against SARS-CoV-2 [2, 3].

The gold standard test for the detection and quantification of neutralizing antibodies is the conventional plaque reduction neutralization test (PRNT). For PRNT, serum is mixed with infectious virus particles and incubated on a cell monolayer so that cytopathic effects (CPE) can be observed. In the case of SARS-CoV-2 this requires a BSL-3 laboratory, restricting PRNT testing to certain laboratories with the appropriate infrastructure. Moreover, the PRNT is time-consuming, with incubation times of 3–5 days in the case of SARS-CoV-2, and more expensive compared to other antibody detection methods such as enzyme-linked immunosorbent assays (ELISA) or lateral flow assays. In order to overcome these disadvantages, alternative methods for the detection and quantification of neutralizing antibodies against SARS-CoV-2 have been developed, including neutralization tests using pseudoviruses, which can be handled in a BSL-2 laboratory [4, 5], and surrogate virus neutralization tests (sVNT) in ELISA format. In contrast to PRNT and pseudovirus neutralization tests, sVNTs produce results within 2–3 hours as opposed to several days.

The first sVNT to detect neutralizing antibodies to SARS-CoV-2 was commercialized by GenScript Biotech in mid 2020 (Piscataway Township, New Jersey, USA) [6] and has obtained FDA approval. In principle, the GenScript sVNT detects antibodies that block the interaction of SARS-CoV-2 with its entry receptor angiotensin-converting enzyme 2 (ACE2). The ACE2 receptor is recognized by the receptor-binding domain (RBD) of the S1 subdomain of the viral spike protein (S-protein). This sVNT has been used, for example, to analyze the longevity of neutralizing antibodies up to day 180 post infection [7] or to identify plasma donors with high titers for antibody therapy [8, 9]. Several similar sVNTs have been developed by different companies (e.g. Euroimmun AG and Beijing Wantai Biological), but to date little independent performance data is available for these tests.

In the present study, we aim to evaluate whether the GenScript sVNT can substitute the PRNT and be used to determine the presence of neutralizing antibodies in vaccinated and convalescent individuals. For this purpose, we analyzed the diagnostic performance of sVNT against PRNT as well as the correlation of antibody levels by using the two methods on a large sample set of about 500 sera. Additionally, we compared the sVNT to three commercial ELISAs (Euroimmun S1 IgG ELISA, Euroimmun NCP IgG ELISA and Wantai complete Ab ELISA) and two other commercial sVNT ELISAs for the specific detection of neutralizing antibodies against SARS-CoV-2 (Wantai SARS-CoV-2 NAbs ELISA and Euroimmun NeutraLISA). Finally, we compared the performance of the GenScript sVNT in sera from convalescent individuals to that in sera from fully vaccinated individuals.

## Materials & Methods

### Sample material

A total of 491 serum samples was collected in the course of routine diagnostics at the Robert Koch Institute, Berlin, Germany. These samples were collected before the end of April 2021, i.e. prior to the large-scale availability of SARS-CoV-2 vaccines to the general German population. In addition, samples from 112 individuals fully vaccinated with different SARS-CoV-2 vaccines (N=74 2x Comirnaty [Biontech], N=14 2x Spikevax [Moderna], N=24 Vaxzevria [AstraZeneca] + Comirnaty) were collected through routine diagnostics. The study obtained ethical approval by the Berliner Ärztekammer (Berlin Chamber of Physicians, Eth 20/40).

### Conventional plaque reduction neutralization test (PRNT)

Briefly, 100 μL of serum were diluted in DMEM (10 % FCS, 2 mM L-glutamine) in six two-fold dilutions, resulting in final serum dilutions of 1:10 up to 1:320. For some samples, 200 μL of serum was used as starting material for two-fold dilutions, leading to final serum dilutions of 1:5 up to 1:160. Dilutions were mixed 1:1 with SARS-CoV-2 (strain BetaCoV/Germany/BavPat1/2020, Institute for Microbiology of the German Armed Forces; final virus concentration 1,000 TCID_50_/mL) and incubated at room temperature (RT) for 1 h. Next, 100 μL of diluted serum–virus mix were added to wells containing 2 × 10^4^ Vero E6 cells per well (#85020206, European Collection of Authenticated Cell Cultures (ECACC), Porton Down, UK) in a 96-well plate. Each sample dilution was tested in eight replicates and cells were incubated for 5 days at 37 °C, 5 % CO_2_. After 5 days each well was analyzed by light microscopy for visible CPE. The number of wells without CPE (negative wells) was counted and PRNT_50_ values were calculated according to Reed and Muench [10]. For quality control, a positive control with defined titer was analyzed in parallel and back-titration of the virus stock was performed. Samples with a titer ≥ 1:15 were considered positive.

### GenScript surrogate virus neutralization test (sVNT)

The GenScript cPass™ SARS-CoV-2 Neutralization Antibody Detection Kit (Genscript Biotech, Leiden, The Netherlands) is based on a two-step process: firstly, the recombinant HRP-coupled RBD is pre-incubated with serum, leading to binding of RBD by present RBD-specific antibodies; and secondly, addition of the mixture to the ELISA plate where antibody-bound RBD and any unbound RBD will compete for binding to the coated ACE2 receptor. Neutralizing antibodies are considered to block the binding between RBD and ACE2, hence increasing the amount of free RBD that can bind to the plate. The raw OD signal is thus inversely correlated to the amount of neutralizing antibodies. Using the negative control as reference, the OD is then converted to a percent value, reflecting the degree of signal inhibition by neutralizing antibodies.

The GenScript sVNT was performed according to the manufacturer’s instructions. Briefly, serum samples as well as negative and positive controls were diluted 1:10 in sample dilution buffer, mixed 1:1 with HRP-RBD working solution and incubated at 37 °C for 30 min. Subsequently, 100 μL of sample and controls were added into the wells of the 96-well plate and coated with the ACE2 receptor. The plate was incubated at 37 °C for 15 min and washed 4x with 300 μL of washing buffer. Next, 100 μL of substrate solution were added and the plate was incubated in the dark for 15 min at RT. Finally, 50 μL of stop solution per well were added and the absorption at 450 nm was measured by using a Tecan infinite M200 Pro ELISA-Reader. The percentage of signal inhibition in relation to the negative control was calculated as follows: Inhibition [%] = (1 – (Sample OD_450_ / Average Negative Control OD_450_)) x 100. All samples and controls were analyzed in duplicate. The analysis was repeated if the coefficient of variation between both replicates was above 10 %.

### Additional ELISA tests to detect SARS-CoV-2-specific neutralizing antibodies

The Wantai SARS-CoV-2 nABs ELISA (Beijing Wantai Biological Pharmacy Enterprise; Beijing, China) for the detection of neutralizing antibodies against SARS-CoV-2 was used according to the manufacturer’s instructions. Briefly, 50 μL of positive and negative control and 100 μl of serum sample were added per well and the plate was incubated at 37 °C for 60 min. Next, the plate was washed 4x with the supplied wash buffer and 100 μL of Spike-RBD antibodies conjugated to HRP were added per well. After incubation at 37 °C for 30 min the plate was again washed 5x, followed by addition of 50 μL of Chromogen Solution A and 50 μl of Chromogen Solution B per well. The plate was incubated at 37 °C for 15 min in the dark and 50 μL of stop solution were added. The absorption at 450 nm was measured by using a Tecan infinite M200 Pro ELISA-Reader and a reference wavelength of 620 nm. Samples were analyzed in single replicate.

The Euroimmun SARS-CoV-2 NeutraLISA (Euroimmun AG, Lübeck, Germany) was performed according to the manufacturer’s instructions. Serum samples were diluted 1:5 in sample buffer and 100 μl of diluted sample, control or blank were added per well and incubated at 37 °C for 1 h. The plate was washed with the supplied wash buffer and 100 μl of enzyme conjugate were added and incubated at RT for 30 min. After another wash cycle 100 μl of substrate solution were added per well and the plate incubated at RT for 15 min. Finally, 100 μl of stop solution were added per well and the absorption at 450 nm was measured by using a Tecan infinite M200 Pro ELISA-Reader and a reference wavelength of 620 nm. Samples were analyzed in single replicate. The inhibition in percent was calculated as follows: 100% - ((extinction sample x 100%)/average extinction blank).

### Binding antibody ELISA tests

The Euroimmun SARS-CoV-2 IgG antibody ELISA (“S1 IgG”), Euroimmun SARS-CoV-2-NCP IgG ELISA (“NCP IgG”, both Euroimmun AG, Lübeck, Germany) and the Wantai SARS-CoV-2 Ab ELISA (Beijing Wantai Biological Pharmacy Enterprise; Beijing, China) were carried out according to the manufacturer’s instructions, with the exception that 50 μl of serum were used in the Wantai Ab ELISA. Cutoffs were used as recommended by the manufacturer (EI ELISAs: ratio <0.8 negative, ≥0.8 to <1.1 equivocal, ≥1.1 positive; Wantai ELISA: ratio <0.9 negative, ≥0.9 to <1.1 equivocal, ≥1.1 positive). Samples were analyzed in duplicate by using the Euroimmun ELISAs and single values were measured by using the Wantai Ab ELISA. In contrast to all other ELISAs, samples for the Euroimmun ELISAs were heat-inactivated at 57 °C for 1 h.

## Results

### Performance of GenScript sVNT as a surrogate test for PRNT

In order to assess the performance of the GenScript sVNT against the gold standard PRNT, we analyzed 491 pre-vaccination sera including 245 PRNT negatives and 246 PRNT positives. Using the manufacturer-recommended positivity cutoff of 30 % binding inhibition, the test showed a sensitivity of 76.4 % and a specificity of 95.9 %. However, a substantial number (58; 23.6%) of PRNT-positive samples were falsely identified as negative by sVNT (**Figure 1A+B**). We therefore assessed the effect of defining different equivocal areas on the test sensitivity and specificity (**Supplementary Table and Figure S1**). Defining a equivocal area between 15 and 35 % binding inhibition resulted in the best sensitivity and specificity which ranged between 72.8–93.1 % and 73.5–97.6 %, respectively, depending on whether indeterminate samples were counted as positive or negative. In this equivocal area, 46.8 % of samples were positive in PRNT with generally low neutralizing antibody titers (median 1:20, IQR: 17–26). Hence, for further analyses we defined in-house cutoffs as follows: <15 % binding inhibition = negative, ≥15 % to <35 % binding inhibition = equivocal and ≥35 % binding inhibition = positive.

**Figure 1.**
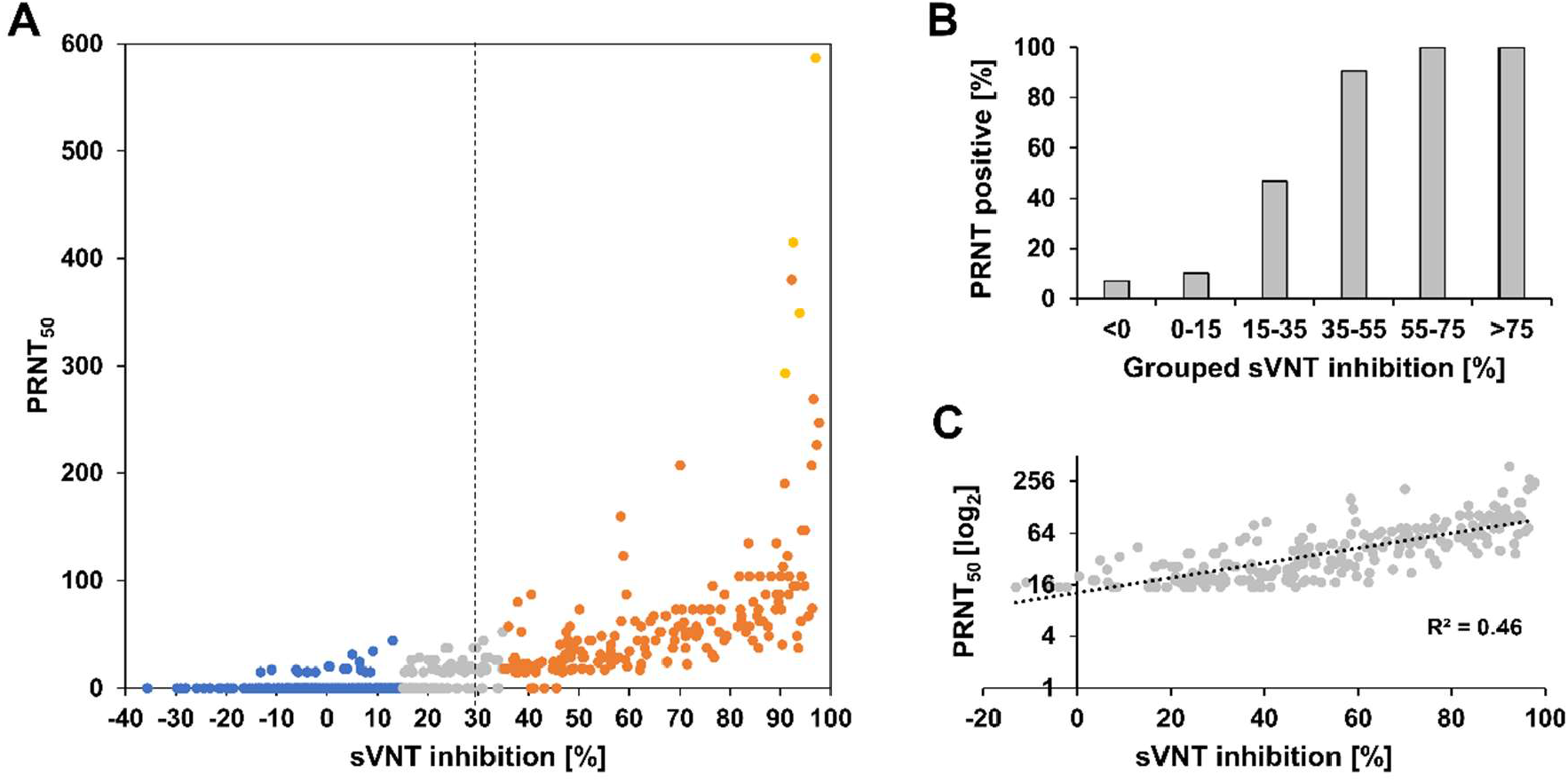
Comparison of GenScript sVNT to PRNT in pre-vaccination sera. (A) Comparison of PRNT (PRNT_50_) and sVNT results (N = 491). Dashed line: manufacturer-recommended positivity cutoff. Data points are color-coded according to the GenScript sVNT result and in-house cutoffs: blue (<15 % = negative), gray (≥15 to <35 % = equivocal) and orange (≥35 % = positive). Samples with titers above the PRNT quantification limit (>) are marked in yellow. (B) Percentage of PRNT-positive samples across grouped sVNT results (N = 491). (C) Correlation of PRNT titer (PRNT_50_) in PRNT-positive samples with the sVNT result (n = 242).

In order to evaluate whether the GenScript sVNT, which is marketed as a semiquantitative assay, can be used to estimate the titer of neutralizing antibodies, we investigated the correlation between PRNT_50_ values obtained from PRNT and binding inhibition values obtained from GenScript sVNT. For PRNT-positive samples, results from the two tests showed a moderate correlation (R^2^=0.60) (**Figure 1C**).

After assessing the performance of the GenScript sVNT ELISA against the gold standard of PRNT, we next aimed to evaluate whether this test is indeed a better surrogate test for PRNT as compared to other common commercial ELISA tests which are not designed to specifically detect neutralizing antibodies. We therefore assessed the performance of the Euroimmun S1 IgG ELISA, Euroimmun NCP IgG ELISA and Wantai complete Ab ELISA in comparison to PRNT, using varying subsets of the 491 samples used for the validation of GenScript sVNT against PRNT. For direct comparability, the correlation of the PRNT titer with Euroimmun or Wantai ELISA and with the sVNT results in the respective subset was analyzed in parallel (**Figure 2A-C**). An overview of the assays and sample subsets used in the present study can be found in **Supplementary table S2**.

**Figure 2.**
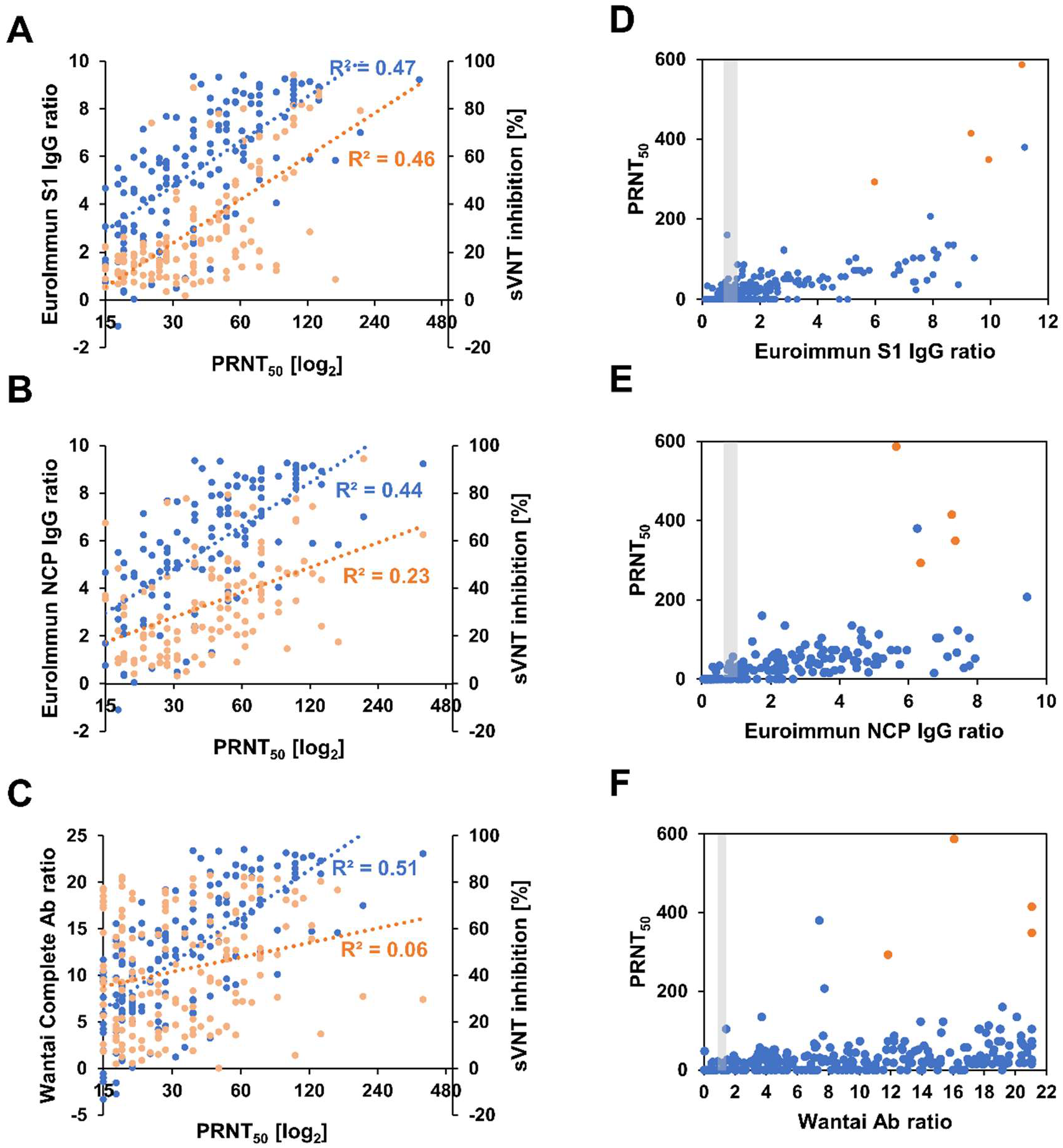
Assessment of further commercial ELISAs as predictive tests for neutralizing antibodies. Correlation of PRNT titer (PRNT_50_) in PRNT-positive samples in the respective sample subsets with (A) the Euroimmun S1 IgG ELISA (light orange) and sVNT results (blue, N =133), (B) the Euroimmun NCP IgG ELISA (light orange) and sVNT results (blue, N =116), (C) the Wantai complete Ab ELISA (light orange) and sVNT results (blue, N =167). Comparison of PRNT results, including PRNT-negative and -positive samples, with (D) the Euroimmun S1 IgG ELISA (N=298), (E) the Euroimmun NCP IgG ELISA (N=205) and (F) the Wantai complete Ab ELISA (N=388). (D-F) Samples shown in orange could not be fully quantified in PRNT as their titer was above the quantifiable limit; the minimal titer estimate is displayed for these samples. Gray-shaded box: equivocal area of the respective ELISA.

As expected, while the sensitivity of the commercial ELISAs against PRNT was comparable to that of the GenScript sVNT, their specificity for neutralizing antibodies was generally lower with positive signals observed in PRNT-negative samples (**Figure 2D-F, Supplementary Table S3**). In PRNT-positive samples, the Euroimmun S1 IgG ELISA and GenScript sVNT results correlated equally well with PRNT titers (**Figure 2A**). As a result, when only samples with a relatively high ratio of >4 in the Euroimmun S1 IgG ELISA were considered, 40 out of 42 (95.2%) were PRNT positive (**Figure 2D**). Euroimmun NCP IgG ELISA results correlated less well with PRNT titers compared the Genscript sVNT results in the respective sample subset (**Figure 2B**), and no correlation was found between Wantai complete Ab ELISA results and PRNT titers (**Figure 2C**). While all 44 samples with a strong signal (ratio >4) in the Euroimmun NCP IgG ELISA were PRNT positive, for the Wantai complete Ab ELISA PRNT-negative samples were observed throughout the dynamic range of the assay (**Figure 2E+F**).

### Performance of GenScript sVNT ELISA on convalescent patients versus vaccinees

One difference in the antibody responses elicited through natural SARS-CoV-2 infections versus vaccination is the breadth of antibody targets. While in natural infections antibodies can be formed against all virus epitopes, vaccination triggers the development of only S-protein-targeting antibodies. We therefore investigated whether the performance of GenScript sVNT, which targets the RBD subunit of the S-protein, differs in samples collected from convalescent patients versus fully vaccinated individuals when compared to PRNT.

Neutralizing antibodies were found by PRNT and sVNT in 104 of 112 (92.9%) vaccinated individuals. Among the nine PRNT-negative samples, two were negative in sVNT, four were indeterminate and three were positive (**Figure 3A**). Binding inhibition values in most positive vaccinees were at the upper limit of quantification of GenScript sVNT, and hence the correlation with PRNT_50_ values could not be investigated formally (**Figure 3B**). In line with this, at similar PRNT_50_, binding inhibition values in vaccinees were always at the upper end of binding inhibition values observed in convalescents (**Figure 3B**). Binding inhibition values were significantly higher in vaccinees (median 95.7, IQR 88.1–96.8) compared to convalescent patients (median 52.9, IQR 31.2–76.2, p<0.001) (**Figure 3C)**. On average, also PRNT_50_ titers were higher in vaccinated individuals (median 119.8, IQR 56.7–160) compared to convalescent patients (median 49.1, IQR 20–62, p<0.001) (**Figure 3D**) but only rarely reached the upper limit of quantification of PRNT. It should be noted that the period between vaccination and sampling in vaccinated individuals was longer (median 10.1 weeks, IQR 6.9–14.7) compared to the period between symptom onset and sampling for convalescent individuals (median 5.1 weeks, IQR 4.3–6.6, p<0.001, **Figure 3E**).

**Figure 3.**
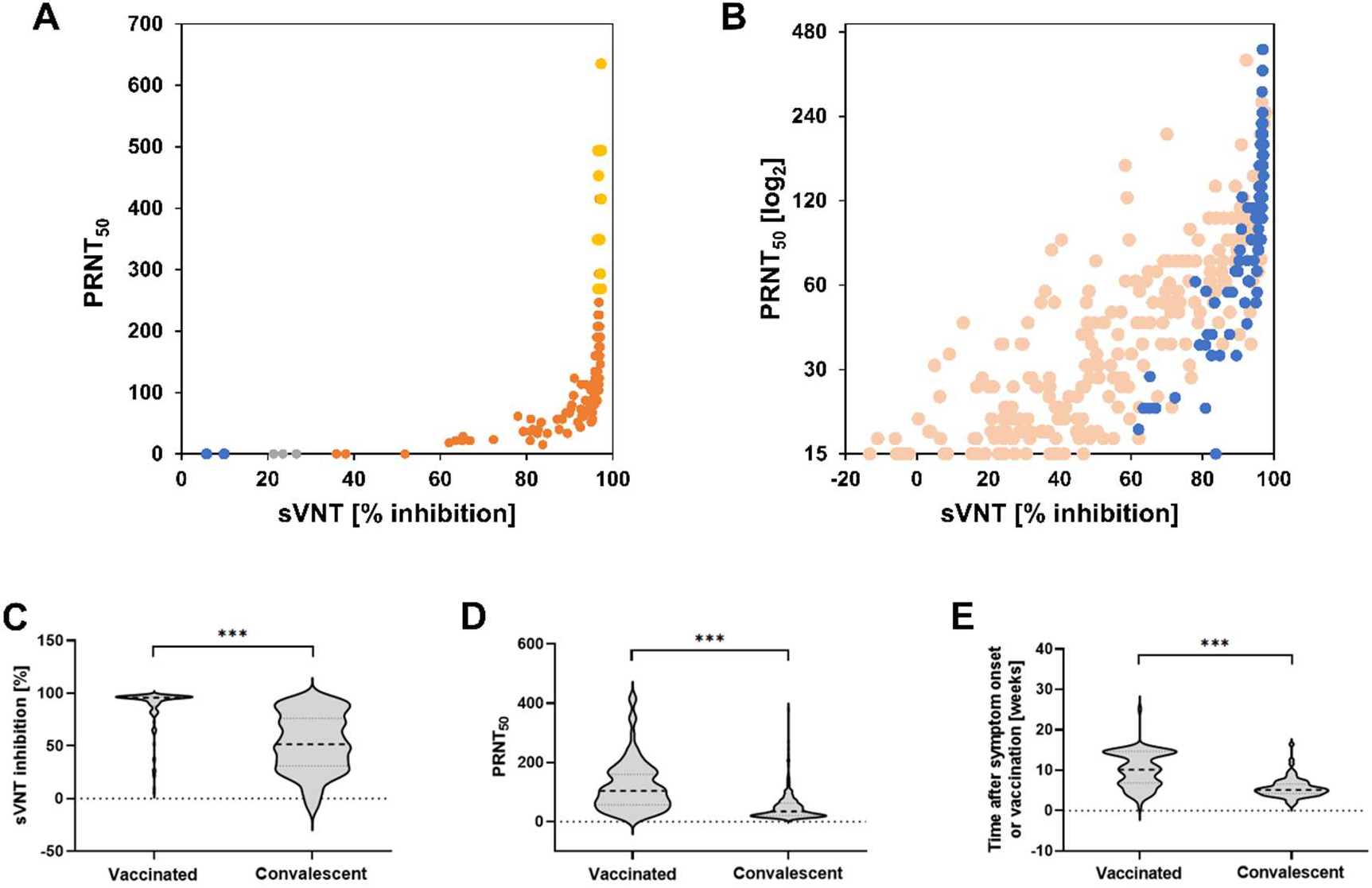
Performance of the sVNT ELISA in fully vaccinated individuals in comparison to convalescent patients. (A) Comparison of PRNT_50_ and binding inhibition in vaccinated individuals (N = 112). Data points are color-coded according to the GenScript sVNT results and in-house cutoffs: blue (<15 % = negative), gray (≥15 to <35 % = equivocal) and orange (≥35 % = positive). Samples with titers above the PRNT quantification limit (>) are marked in yellow. (B) Comparison between sVNT and PRNT (PRNT_50_) in vaccinated (blue; N = 90) versus convalescent (light orange; N = 242; data from Figure 1C) individuals, including only PRNT-positive samples. (C) Distribution of the sVNT-binding inhibition results in vaccinated (N = 112) and convalescent individuals (N = 242). (D) Distribution of PRNT_50_ titers in vaccinated (N = 90) and convalescent (N = 242) individuals. (E) Time difference between symptom onset or vaccination (N = 90) and convalescent (N = 75) individuals for whom the period between symptom onset or vaccination and sampling was known. Statistics: t-test, ***p≤0.001.

### Performance of alternative surrogate ELISA tests against PRNT

Finally, we investigated the agreement between different surrogate ELISAs that are marketed specifically for the detection of neutralizing antibodies against SARS-CoV-2. In addition to the GenScript sVNT, we tested the Euroimmun NeutraLISA and Wantai NAbs ELISA. In these tests, neutralizing antibodies in the serum compete either with the ACE2 receptor for binding to coated RBD protein (NeutraLISA) or with anti-RBD antibodies for binding to coated S antigen (Wantai Nabs ELISA).

Agreement in positivity between the GenScript sVNT and the Wantai NAbs ELISAs was moderate to strong in a subset of samples used for the validation of the GenScript sVNT (N=111; GenScript sVNT indeterminate samples considered as positive, Cohen’s kappa: 86.6%; considered as negative: 70.4%). However, 8 PRNT-negative samples that were classified as indeterminate by the GenScript sVNT were falsely classified as positive by using the Wantai NAbs ELISA (**Figure 4A**) since no equivocal area is the defined for this ELISA by the manufacturer. Despite a good sensitivity against PRNT (98.7%), the specificity was thus in the lower range (75.8%) of that observed when using the GenScript sVNT in this sample set (sensitivity 89.7–100%; specificity 63.6–97.0%, depending on whether indeterminate samples were counted as positive or negative). Agreement in positivity between the GenScript sVNT and the Euroimmun NeutraLISA was weak at best (indeterminate samples considered as positive, Cohen’s kappa: 35.2%; considered as negative: 46.8%) in the same subset of samples. Although the correlation between the two tests was good (R^2^ = 0.8, **Figure 4B**), the manufacturer-recommended cutoffs for NeutraLISA were not ideal in our sample set. The equivocal area defined by the manufacturer contained only PRNT-positive samples and 25/78 (32%) of PRNT-positive samples were classified as negative. Hence, despite a perfect specificity against PRNT (100%), the sensitivity of the NeutraLISA was low (50.0–67.9%).

**Figure 4.**
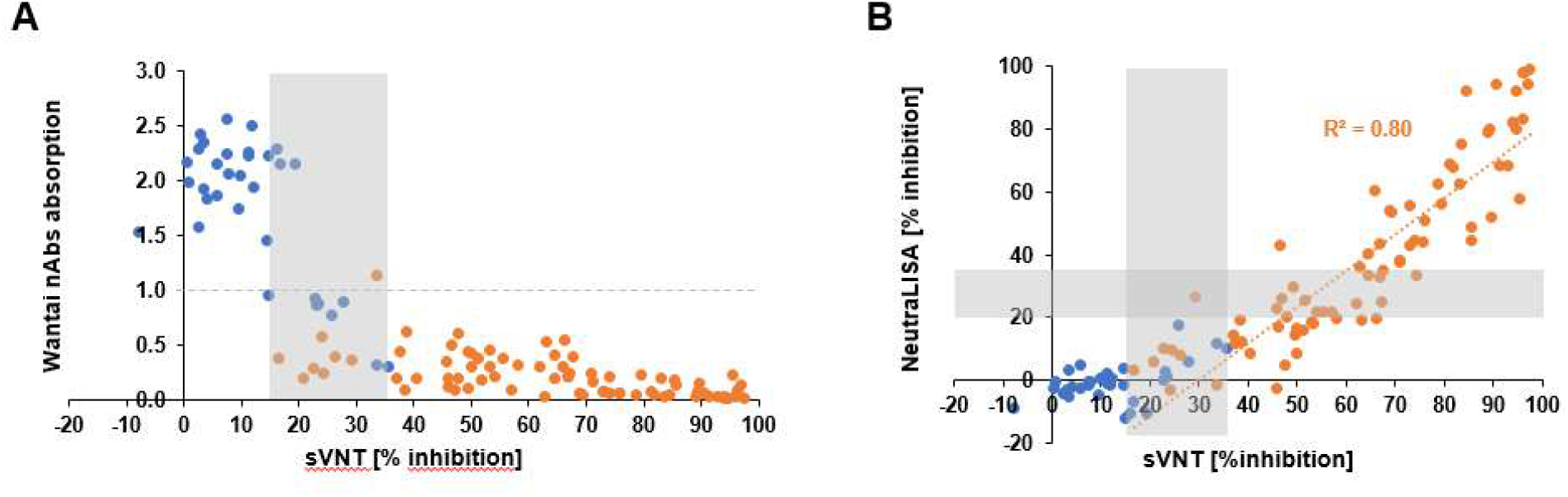
Comparison of Genscript sVNT with the Wantai SARS-CoV-2 NAbs ELISA (A) and the Euroimmun NeutraLISA (B). Data points are color-coded according to their positivity in PRNT (blue: PRNT negative, orange: PRNT positive). Gray dashed line in (A): Wantai nABs ELISA manufacturer-recommended cutoff. Shaded areas in (A) and (B): equivocal areas; in-house for GenScript sVNT and manufacturer-recommended for Euroimmun NeutraLISA. (A) As the Wantai Nabs ELISA is marketed for qualitative use, the manufacturer does not recommend a conversion of the raw data into binding inhibition and hence no correlation to the sVNT was calculated. Due to the competitive nature of the assay, the signal is inversely correlated with the amount of neutralizing antibodies in a sample. Identical sample sets were used in A and B (N = 111).

## Discussion

Surrogate virus neutralization tests, such as the GenScript sVNT ELISA, have the potential to allow a faster detection and quantification of neutralizing antibodies in serum samples when compared to traditional cell culture-based methods such as PRNT and can be used in laboratories without BSL-3 facilities. In addition, as a standardized commercial test, GenScript sVNT results can be considered as better comparable between laboratories than antibody titers obtained by PRNT, where the measured titers depend on the experimental setup (e.g. number of virus particles used, virus strain, cells and serum dilution steps) and can only be compared between laboratories with caution. As the sVNT procedure is performed in plate format and can be either fully or partially automated, also sample throughput is substantially increased. Given the need for rapid detection of neutralizing antibodies in large sample numbers in vaccine trials, for investigating population immunity during the course of the COVID-pandemic or for ascertaining individual vaccination success, access to a rapid and accurate test for detecting and possibly even quantifying neutralizing antibodies to SARS-CoV-2 is crucial.

Previous studies have described a moderate to good correlation of GenScript sVNT results with antibody titers measured by PRNT [6, 11-13]. However, in some of these studies the correlation with PRNT has been assessed by using IC_50_ values determined by sVNT rather than by using the direct signal output of the GenScript sVNT, which is defined as percent of inhibition. While this approach still allows a faster detection and quantification of neutralizing antibodies as compared to traditional PRNT, the sample throughput remains low. In the present study we therefore used the direct GenScript sVNT results, percent binding inhibition, to evaluate the test performance against PRNT. Moreover, we used the largest dataset known so far, containing 491 PRNT-negative and -positive pre-vaccination serum samples plus 112 samples from fully vaccinated individuals. In our dataset, vaccinated individuals generally exhibited higher binding inhibition and neutralizing antibody titers compared to convalescent individuals. Higher antibody titers in vaccinated compared to convalescent individuals have been detected before [14, 15]. In addition, the design of the GenScript sVNT ELISA could further artificially enhance the difference in binding inhibition between vaccines and convalescent individuals: Only S-targeting neutralizing antibodies are present after vaccination with the current SARS-CoV-2 vaccines. In contrast, neutralizing antibodies that are present after natural infections target also virus epitopes other than the S-protein RBD, including other virus proteins. Hence this could result in an underestimation of neutralizing antibodies in convalescent individuals by the GenScript sVNT (which targets the RBD of the S-protein), and it can be conceived that the GenScript sVNT can more accurately quantify neutralizing antibodies in vaccinees compared to convalescent patients.

Low antibody titers between 1:10 and 1:30 PRNT_50_ have been associated with protection against symptomatic SARS-CoV-2 infection in convalescent patients in one meta-analysis [16], underlining the importance of high sensitivity (at high specificity) of any PRNT surrogate test. It should be noted here that the detection limit of the PRNT performed in this study was 1:15 PRNT_50_, resulting in a potential underestimation of PRNT-positive samples. In this study, the sensitivity and specificity of the GenScript sVNT against PRNT in sera collected before the wide availability of SARS-CoV-2 vaccines in Germany was largely determined by the cutoff values used for analysis. Initially, the manufacturer-recommended positivity cutoff for the GenScript sVNT ELISA was set at 20% binding inhibition, which was later raised to 30% by the manufacturer in order to improve specificity for FDA certification. Using a cutoff of 20% binding inhibition, a study by Mueller und colleagues showed a sensitivity of 98.7 % and a specificity of 69.5 % of the GenScript sVNT ELISA [17]. Also, a study by Valcourt et al. using a 20% cutoff showed a substantial rate (32.4–58.3 %) of false positive detections by sVNT compared to PRNT [11]. Papenburg et al.’s investigation of different cutoff values found that increasing the threshold to 30% resulted in a minimal loss of sensitivity (95.2 to 92.8%) but a much larger gain in specificity (44.8 to 57.7%) across varying sample panels [18]. In a study by Nicholson and colleagues, the authors tested three different positivity cutoffs for the GenScript sVNT (20 %, 20 % with retesting in a equivocal area of 18–22 % and 25 %) and found that both alternatives to the initial manufacturer-recommended cutoff of 20% improved specificity [19]. Through our systematic comparative evaluation of different equivocal areas around these varying cutoffs, using a larger sample set than previously published, we show that roughly half of samples from convalescent individuals with 15–35% binding inhibition are positive in PRNT. Setting a equivocal area between 15 and 35% binding inhibition thus allowed us to increase test sensitivity in convalescent patients from 76.4 to a maximum of 93.1% as compared to using a hard cutoff at 30% binding inhibition. As a result of the generally higher titers in vaccinees, the proportion of indeterminate detections by using our in-house-established cutoffs for the sVNT was lower in vaccinees compared to convalescents. A total of three out of nine PRNT-negative samples with sVNT-binding inhibition values of up to 52% suggest that an adaptation of the equivocal area could be advisable to avoid false positive results when analyzing samples from vaccinated individuals by using the GenScript sVNT. However, the validation of a vaccine-specific cutoff would require a larger dataset with the appropriate metadata, including the type of vaccine and time period between vaccination and sampling, than that available in this study.

As mentioned before, antibody titers by PRNT after a full vaccination were generally higher than those in convalescent patients in our sample sets. Samples from vaccinees were taken later (median 10 weeks after the second vaccine dose) compared to samples from convalescents (median 5 weeks after onset of symptoms), although lack of data on the date of infection for many samples collected from convalescent patients may have introduced a bias. SARS-CoV-2 antibody titers have been shown to decline over the first few months after infection [7]; however, a longer period of avidity maturation could in turn lead to a better neutralization and thus to higher detectable titers in PRNT. Higher antibody titers in vaccinated individuals (two doses of Comirnaty) compared to convalescent individuals have also been found in a study from Israel et al. [15], which also described a more rapid decline in antibody titers in vaccinees compared to convalescent individuals. More studies are thus warranted to further elucidate the interplay between maximum antibody titer, rate of titer decline and avidity in convalescent versus vaccinated individuals.

As expected, when comparing the GenScript sVNT test performance to commercial ELISA tests that do not specifically detect neutralizing antibodies, the specificity of the sVNT was superior to that of the other commercial ELISAs. For the two tested Euroimmun ELISAs, but not the Wantai ELISA, high ratios were predictive of the presence of neutralizing antibodies in the sample. The Euroimmun S1 IgG ELISA was previously shown to have a moderate correlation with PRNT (R^2^=0.46 to 0.57) [13, 20], in line with the data from our study (R^2^=0.46). Furthermore, the Euroimmun S1 IgG ELISA ratios and GenScript sVNT results showed a similar correlation with PRNT titers in our study. Using adapted positivity cutoffs for the Euroimmun S1 IgG ELISA, this assay could hence be a useful pre-screening assay that has already been established in many laboratories, with follow-up confirmatory testing of samples with a low positive ratio by using the GenScript sVNT ELISA. In contrast to the Euroimmun S1 IgG ELISA and GenScript sVNT, correlation between the Euroimmun NCP IgG ELISA and PRNT was poor (R^2^=0.23). This can be explained by the different targets used in both assays. The NCP ELISA uses a recombinant N-protein which is known to be a less frequent target for neutralizing antibodies, as compared to the S-protein RBD which is used in the sVNT and is the main target of neutralizing antibodies [21]. When comparing the Wantai complete Ab ELISA ratios and PRNT titers, we observed a complete lack of correlation which can be explained by three major differences between the Wantai ELISA and the other ELISAs: Firstly, opposed to all other tested ELISAs, which are marketed as semi-quantitative assays, the Wantai Ab ELISA is a purely qualitative assay. Secondly, only the Wantai Ab ELISA detects all antibody classes including IgMs, which less frequently have a neutralizing function compared to avidity-matured IgGs, leading to larger differences in the detected antibody populations to PRNT than with the other ELISAs. Thirdly, a 50-fold (this study) to 100-fold (manufacturer’s recommendation) higher sample volume used in the Wantai Ab ELISA as compared to the other ELISAs results in a large proportion of samples with ratios above the dynamic range of the assay, where further quantification and thus determination of a potential predictive cutoff for presence of neutralizing antibodies is not possible.

Comparing the GenScript sVNT against two more recently available commercial neutralizing antibody ELISAs from Wantai and Euroimmun, the sVNT showed the best diagnostic performance in comparison to the gold standard PRNT. Although the NeutraLISA from Euroimmun showed a specificity of 100 %, the manufacturer-recommended equivocal area was not suitable for our dataset as it contained exclusively PRNT-positive samples and therefore did not meet the criteria of a equivocal area. For the GenScript sVNT we applied our in-house equivocal area, while the Wantai NAbs ELISA employs a hard cutoff which is calculated relative to the negative controls according to the manufacturer’s recommendations. However, the definition of a equivocal area would also be beneficial for the Wantai NAbs ELISA to avoid reporting of false positive results.

There are some technical limitations concerning the use of the GenScript sVNT for high-throughput sample analysis. We have identified pipetting speed as one of the main factors involved in the generation of false positive results, as the strength of the signal is strongly correlated with incubation time. If pipetting occurs slowly (e.g. when using a single-channel pipette), HRP-coupled RBD will have more time to bind to the coated ACE receptor, and will thus create a stronger OD signal, in negative samples at the beginning of the plate compared to negative samples at the end of the plate. In our laboratory, where the GenScript sVNT is performed manually, we were able to address this “OD drift” over the plate by using a multichannel pipette and by processing only half plates at a time. However, for full automation this time-dependent drift can be problematic as many pipetting robots use a single-tip dispenser and, hence, robots often cannot adhere to the 2-min pipetting time per plate as recommended by the manufacturer. Nonetheless, the throughput achievable by using the GenScript sVNT remains much higher than that of a cell-culture-based PRNT, even if only half plates are processed. However, it impairs the large-scale testing of vaccinated individuals on a routine basis.

Another parameter that remains to be evaluated when using the GenScript sVNT is the influence of emerging variants with mutations in the RBD, e.g. B1.1.7, B.1.351 and P1. A possible solution would be the development of adapted versions of the kit with the respective mutated RBDs.

## Conclusion

In conclusion, the GenScript sVNT ELISA is suitable to specifically detect neutralizing antibodies in convalescent and vaccinated individuals. It is a valuable screening assay for neutralizing antibodies against SARS-CoV-2 due to its high throughput and short time-to-result, compared to PRNT. However, to achieve high sensitivity and specificity, a equivocal area around the manufacturer cutoff is recommended. This equivocal area, where follow-up testing by PRNT is required, is especially important to consider when accurate individual results are required. For use in vaccinated individuals, positivity cutoffs may require adaptation, and, due to high signals above the limit of quantification of GenScript sVNT, the quantification of neutralizing antibodies is often not possible when using the standard manufacturer’s protocol.

## Data Availability

All data produced in the present study are available upon reasonable request to the authors

## Acknowledgments

The authors are grateful to the colleagues that supported the serological testing in ZBS 1 and to Ursula Erikli for copy-editing.

