## Supplementary information for "Detection of neutralizing antibodies against SARS-CoV-2 by using a commercial surrogate virus neutralization ELISA: can it substitute the classical neutralization test?"

**Content**

**Table S1.** Evaluation of the effect of different cutoffs and equivocal areas on the diagnostic performance of the GenScript sVNT……………………………………………………………………………………………2

**Table S2.** Details of tested ELISAs and sample subsets used for the comparative evaluation……………..2

**Table S3.** Comparison of the diagnostic performance of GenScript sVNT to other commercial ELISAs.3

**Figure S1.** Receiver operating characteristic (ROC) curve for the GenScript sVNT………………………………3

**Table S1. Evaluation of the effect of different cutoffs and equivocal areas on the diagnostic performance of the GenScript sVNT**

| | | Number of samples (classified by GenScript sVNT) | | | Counting indeter-minate as positive | | Counting indeter-minate as negative | |
|---|---|---|---|---|---|---|---|---|
| | PRNT | Positive | Indeter-minate | Negative | Sensitivity | Specificity | Sensitivity | Specificity |
| No equivocal area (cut-off at 30%) | pos | 188 | 0 | 58 | 76.42% | 95.92% | 76.42% | 95.92% |
| | neg | 10 | 0 | 235 | | | | |
| Equivocal area from 10-40% | pos | 159 | 71 | 16 | 93.50% | 57.96% | 64.63% | 97.55% |
| | neg | 6 | 97 | 142 | | | | |
| Equivocal area from 15-35% | pos | 177 | 52 | 17 | 93.09% | 73.47% | 71.95% | 97.55% |
| | neg | 6 | 59 | 180 | | | | |
| Equivocal area from 20-30% | pos | 188 | 32 | 26 | 89.43% | 87.35% | 76.42% | 95.92% |
| | neg | 10 | 21 | 214 | | | | |

**Table S2. Details of tested ELISAs and sample subsets used for the comparative evaluation.**

| **ELISA** | **Type** | **Qualitative / Quantitative** | **Target** | **No of samples tested in comparison to PRNT** |
|---|---|---|---|---|
| Detection of neutralizing Antibodies | | | | |
| GenScript cPass SARS-CoV-2 Neutralization Ab detection kit | Competitive | Semiquantitative | RBD | 491 convalescent<br>112 vaccinated |
| Wantai NAbs | Competitive | Qualitative | S | 111 |
| EuroImmun SARS-CoV-2 NeutraLISA | Competitive | Semiquantitative | RBD | 111 |
| Detection of binding Antibodies | | | | |
| Euroimmun S1 IgG ELISA | Indirect | Semiquantitative | S1 | 298 |
| Euroimmun NCP IgG ELISA | Indirect | Semiquantitative | NCP | 205 |
| Wantai complete Ab ELISA | Sandwich | Qualitative | RBD | 388 |

**Table S3. Comparison of the diagnostic performance of GenScript sVNT to other commercial ELISAs, using PRNT as gold standard**

| | | **Commercial ELISA** (Euroimmun S1 IgG / Euroimmun NCP IgG / Wantai complete Ab, respectively) | | | | | **GenScript sVNT** (using in-house validated cutoffs) | | | | |
|---|---|---|---|---|---|---|---|---|---|---|---|
| | PRNT | Pos | IND[1] | Neg | Sensitivity[2] | Specificity[2] | Pos | IND[1] | Neg | Sensitivity[2] | Specificity[2] |
| Full sample set | pos | | | | | | 185 | 52 | 17 | 72.8 - 93.3% | 73.8 - 97.6% |
| | neg | | | | | | 6 | 60 | 180 | | |
| Euroimmun S1 IgG sample set | pos | 113 | 14 | 10 | 82.5 - 92.7% | 64.0 – 78.9% | 105 | 23 | 9 | 76.8 - 93.% | 76.4 - 98.8% |
| | neg | 34 | 24 | 103 | | | 2 | 36 | 123 | | |
| Euroimmun NCP IgG sample set | pos | 103 | 12 | 5 | 85.8 - 95.8% | 85.9 - 90.6% | 98 | 13 | 9 | 81.7 - 92.5% | 75.3 - 98.8% |
| | neg | 8 | 4 | 73 | | | 1 | 20 | 64 | | |
| Wantai complete Ab sample set | pos | 178 | 0 | 4 | 97.8% | 71.8% | 122 | 43 | 17 | 67.0 – 90.7% | 74.8 - 97.6% |
| | neg | 58 | 0 | 148 | | | 5 | 47 | 154 | | |

[1] IND: Indeterminate

[2] Ranges depend on whether indeterminate samples were counted as positive or negative

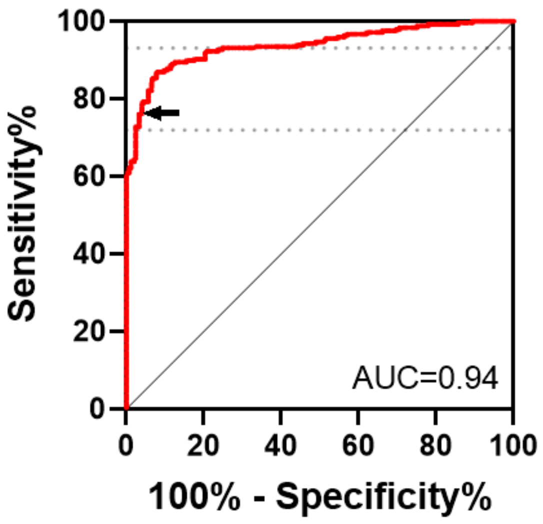


**Figure S1. Receiver operating characteristic (ROC) curve for the GenScript sVNT.**
Arrow: manufacturer-recommended cutoff (30% inhibition). Grey dashed lines: in-house defined equivocal area (15-35% inhibition).
